# Evaluating the Significance of Atypical and Malignant Breast Lesions Arising in Fibroadenoma of the Breast

**DOI:** 10.1101/2022.12.11.22283315

**Authors:** Sunder Sham, Doaa Morrar, Reham Al-Refai, Ahmed Bandari, Azar Garajayev, Sabina Hajiyeva

## Abstract

**Context:** While fibroadenomas (FAs) are the most common benign neoplasm of the breast, little data exists about the clinical characteristics and prognostic value of FAs found to be directly associated with atypical and malignant lesions.

**Objective:** Cases of FA of the breast were reviewed to establish the exact clinical significance of these lesions involved by atypia and malignancy.

**Design:** All FA cases diagnosed on core needle biopsy (CNB) between 2013 and 2022 were screened to identify atypical and malignant lesions {(atypical lobular hyperplasia (ALH), lobular carcinoma in situ (LCIS), atypical ductal hyperplasia (ADH), flat epithelial atypia (FEA), ductal carcinoma in situ

(DCIS), invasive ductal carcinoma (IDC), and invasive lobular carcinoma (ILC)} arising in FA. The relationship between CNB and excisional findings for each case was reviewed.

**Results:** A total of 1500 cases of FA diagnosed on CNB were identified. Among these, 23 cases showed atypical and/or malignant lesions within FA. The median age at diagnosis was 53 years. Of those 23 cases, five were LCIS, one was LCIS+ILC, one was LCIS+ALH, one DCIS+IDC, one was LCIS+DCIS, three were ADH, seven were ALH, and four were DCIS. For LCIS, the excision showed LCIS (2/5), LCIS + ALH (1/5), LCIS + DCIS (1/5), LCIS + DCIS + ALH (1/5). For DCIS, excisions showed Invasive carcinoma with mixed ductal and lobular features (1/4), LCIS + DCIS (1/4), LCIS + DCIS + IDC [classic and pleomorphic type] (1/4), no residual carcinoma (1/4). For ALH, excision showed ALH (2/7), no residual ALH (2/7), ALH+IDP (Intraductal Papilloma) (2/7), LCIS (1/7). For ADH, DCIS (1/3), and benign findings (2/3). For LCIS+ILC and DCIS+IDC, the excisional findings were the same. For LCIS+ ALH, the excisional findings showed benign findings with the radial scar (1/1).

**Conclusion:** There is a low percentage of FA harboring atypia or carcinoma. Due to the high upgrade rate following excision, complete excision of these lesions may guide recommended method of clinical management

## Introduction

Fibroadenomas (FAs) are the breast’s most common benign epithelial neoplasm, accounting for 50% of all breast biopsies.^1^ FAs usually present with a well-defined, mobile mass on physical examination or a solid mass on ultrasonography, and a definitive diagnosis can be made with a core needle biopsy (CNB) or excision.^1^ FAs are characterized by an admixture of stromal and epithelial tissue. They arise from the breast’s lobules (milk-producing glands) and are generally painless, rubbery, and mobile. On CNB, they consist of abundant stromal cells.^1^

Benign epithelial breast lesions can be classified based upon cellular proliferation and atypia degree into nonproliferative, proliferative without atypia, and atypical hyperplasia.^2^ While most simple FAs have not shown an increased risk of breast cancer, other specific benign breast lesions, e.g., a proliferative disease or those with a family history of breast cancer, have a slightly increased risk. The chance of carcinoma, however, arising in a fibroadenoma is meager (0.1-0.3%), and only about 100 cases have been reported in the medical literature.^3^ They usually are incidental pathologic findings on CNB or after excision of the FA. Most reported cases show in situ malignancy; however, invasive breast carcinoma can develop in FA.^4^ Patients with proliferative breast lesions with atypia, including atypical ductal hyperplasia (ADH), atypical lobular hyperplasia (ALH), and lobular carcinoma in situ (LCIS), are at increased risk of developing breast cancer. On excisional biopsy, the percentage of an upgrade of ADH, ALH, and LCIS into ductal carcinoma in situ (DCIS) and invasive breast cancer is 10-20%, <3%, and <3%, respectively.^5,6^

While FAs are the most common benign neoplasm of the breast, little data exists about clinical risks, characteristics, and prognostic value of atypical and malignant lesions arising within FAs. For example, invasive ductal carcinoma (IDC) in FA is sporadic, with an incidence of 0.02%–0.125%.^7,8^ Although rare but it carries the risk of malignancy. The data on the malignant potential of FA is scarce. There are no definite clinical or radiological criteria for diagnosing carcinoma developing in a fibroadenoma.

We reviewed the cases of FA diagnosed within our health system to establish the rate and clinical significance of FAs with atypia and/or carcinoma. In addition, the current study also aimed to assess the rate of atypia and carcinoma diagnosed on CNB material and compare it to the findings in subsequent excisional specimens.

## Materials and Methods

A retrospective review was conducted to identify all FA cases diagnosed on core CNB between 2013 and 2022 by searching the pathology database within our pathology department. All FA cases were then screened to analyze those with atypical and malignant lesions [ALH, LCIS, ADH, FEA, DCIS, IDC, and ILC] arising in FA. The total sample size was 1500.

### Inclusion criteria

All patients where atypia, in situ, or invasive carcinoma were limited to fibroadenoma on CNB were included. *Exclusion criteria:* Cases with atypical or malignant lesions outside of fibroadenoma on the same CNB and those with a previous or current history of breast malignancy were excluded; all cases that did not have subsequent excisional material for our review were also excluded from the study.

All fibroadenoma cases identified to have atypical/malignant lesions arising within the lesion were analyzed, reviewed, and compared through biopsy, excision, and final diagnosis, as depicted in figure 1.

**Figure 1:**
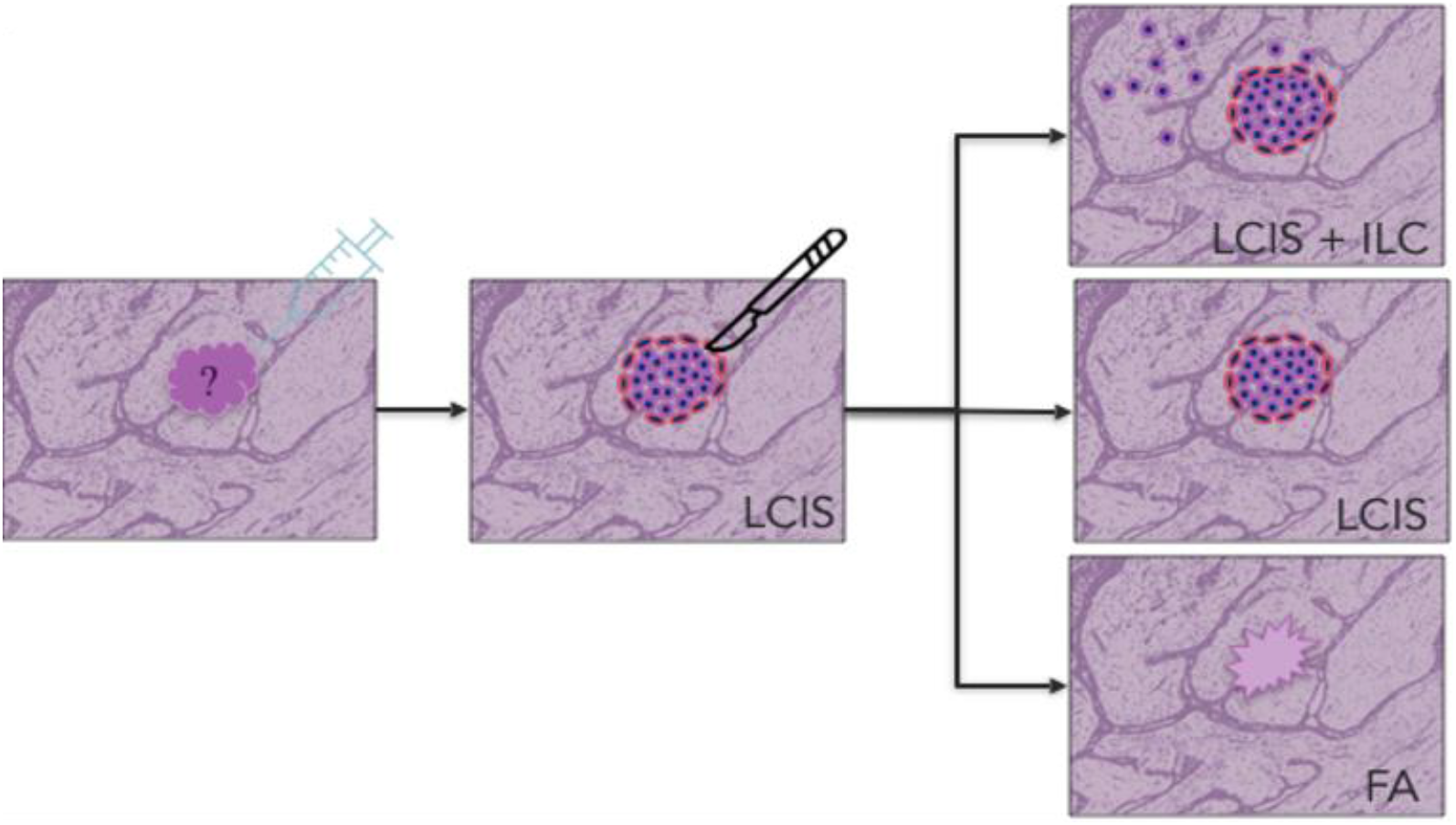
**All fibroadenoma cases identified as atypical/malignant lesions arising within the lesion were analyzed through biopsy, excision, and final diagnosis. Example cases identified LCIS within FA on CNB and subsequent findings in excisional material**.

### Statistical Analysis

Data was analyzed using categorical/qualitative variables such as gender and biopsy features of malignancy, rate of atypia or carcinoma was calculated as frequency and percentage. Ethical Approval: All human participants’ procedures were approved by the Institutional Review Board of Lenox Hill Hospital, Northwell Health, New York, USA.

## Results

A total of 1500 cases of FA diagnosed on CNB were identified. Among these, 23 cases showed atypical and/or malignant lesions within FA (0.01%). The median age of the patients at diagnosis was 53 years (range: 40-68 years). Of those 23 cases, Of those 23 cases, five were LCIS, one was LCIS+ILC, one was LCIS+ALH, one DCIS+IDC, one was LCIS+DCIS, three were ADH, seven were ALH, and four were DCIS.For LCIS, the excision showed LCIS (2/23), LCIS + ALH (1/23), LCIS + DCIS (1/23), LCIS + DCIS + ALH (1/23). For LCIS, the excision showed LCIS (2/5), LCIS + ALH (1/5), LCIS + DCIS (1/5), LCIS + DCIS + ALH (1/5). For DCIS, excisions showed Invasive carcinoma with mixed ductal and lobular features (1/4), LCIS + DCIS (1/4), LCIS + DCIS + IDC [classic and pleomorphic type] (1/4), no residual carcinoma (1/4). For ALH, excision showed ALH (2/7), no residual ALH (2/7), ALH+IDP (Intraductal Papilloma) (2/7), LCIS (1/7). For ADH, DCIS (1/3), and benign findings (2/3). For LCIS+ILC and DCIS+IDC, the excisional findings were the same. For LCIS+ ALH, the excisional findings showed benign findings with the radial scar (1/1). (Table 1)

**Table 1:** All fibroadenoma cases with atypical/malignant lesions are numerically and in percent shown both on Core-Needle (CNB) and Excisional Biopsy.

| **CNB Diagnosis (n/%)** | **Excisional Diagnosis (n/%)** |
| --- | --- |
| LCIS (5/23) -22% | ● LCIS (2/5) – 40%<br>● LCIS + ALH (1/5) – 20%<br>● LCIS + DCIS (1/5) -20%<br>● LCIS + DCIS + ALH (1/5) – 20% |
| DCIS (4/23) – 17.4% | ● Invasive carcinoma with mixed ductal and lobular features (1/4) – 25%<br>● LCIS + DCIS (1/4) -25%<br>● LCIS + DCIS + IDC [classic and pleomorphic type] (1/4) – 25%<br>● No residual carcinoma (1/4) – 25% |
| ALH (7/23) - 30.5% | ● ALH (2/7)– 28.6%<br>● No residual ALH (2/7) -28.6%<br>● ALH+IDP (2/7) – 28.6%<br>● LCIS (1/7) – 14.2% |
| ADH (3/23) -13.1% | ● DCIS (1/3) – 33.3%<br>● Benign findings (2/3) - 66.7% |
| LCIS and ILC (1/23) - 4.5% | ● LCIS + DCIS (1) -100% |
| DCIS and IDC (2/23) - 9% | ● LCIS + DCIS (2/2) -100% |
| LCIS and ALH (1/23) – 4.5% | • Benign findings [radial scars] (1/1) – 100% |

## Discussion

FAs are the most common tumor in the breast and are not usually considered a risk factor for developing breast cancer. The chance of carcinoma arising in a fibroadenoma is scarce (0.1-0.3%), and only about 100 cases have been reported in medical literature to date.^3^ A previous study reported only 5 cases among 4000 FA cases whereas another study showed that among 302 breast carcinoma patients, only 4 cases originated from FAs.^9^ In the current study, we reported an extremely low rate (0.01%) of breast carcinoma arising from fibroadenoma. Among these 23/1500 cases, the mean age of the patients at diagnosis was 53 years, which is older than patients diagnosed with benign FAs (ranging between 20 – 30 years).^10^ In our collective case analysis, the major histologic type (based on CNB) of breast carcinoma arising from FAs were LCIS (22%), LCIS + ILC (4.5%), DCIS (17.4%), ALH (30.4%), ADH (13.1%) and DCIS + IDC (9%). These findings are consistent with previous reports that carcinomas arising within FAs are mainly carcinoma in situ (LCIS 67%, DCIS 12.5%) and invasive carcinoma (IDC 11%, ILC 3.4%).^10^ Another study also reported that lobular carcinoma constitutes a higher proportion of carcinomas arising in FAs, and 81% were reportedly in situ carcinomas.^9,11^ Interestingly, only 10% of all breast carcinomas constitute lobular carcinomas in the general population which is s lower rate than the reported rate of lobular carcinomas arising from FAs. The underlying reason for this association might be because lobular carcinoma is both multicentric bilateral and fibroadenomatous epithelial components originate from intralobular or terminal ducts, thus could be the origin of lobular carcinoma. Our current study is a collective analysis of the highest number of cases from FAs till date.

FAs are often suspected clinically and diagnosed on radiological studies; however, histopathologic examination need to confirm benign nature of the diagnosis. Thus, radiologists and sonologists should be cognizant of any focal morphologic changes in an originally benign lesion. Irregular borders and pleomorphic and linear calcifications on a mammogram may raise suspicion for malignancy, but unfortunately, FAs harboring carcinoma cannot often be reliably distinguished from benign FAs via routine imaging. Thus, only CNB and/or excisional biopsy can lead the confirmatory diagnosis.

In the LCIS-harboring tumors diagnosed on CNB, excisional biopsy revealed LCIS (40%, 2/5), LCIS+ALH (20%, 1/5), both LCIS and DCIS (20%, 1/5), and both LCIS, DCIS, ALH (20%, 1/5). In ADH-harboring tumors, excisional biopsy revealed DCIS (33.3%, 1/3) and benign findings (66.7%, 2/3). In DCIS-harboring tumors, excisional biopsy revealed invasive carcinoma with mixed ductal and lobular features (25%, 1/4) and both DCIS, LCIS and IDC (25%, 1/4), LCIS+DCIS (25%, 1/4) and no residual carcinoma (25%, 1/4). However, both CNB and excision showed same lesions for the LCIS + ILC, and IDC + DCIS cases. The tumors that we report in our study were those limited to FA or in the surrounding breast tissue only as confirmed after excision i.e., lumpectomy.

The etiology of FAs is not known, but a hormonal relationship is likely since they persist during the reproductive years, can increase in side during pregnancy or with estrogen therapy, and usually regress during menopause. Understanding the mechanism of malignant transformation in FAs of the breast is critical for a better understanding of the histopathology of the tumor. In most studies menopause and previous pregnancies have been attributed to this transformation. However, Goldenberg et al. and Brown et al. reported a possible association with exogenous hormones (e.g., oral contraceptives).^12,13^ In the literature, two different malignant transformation has been noted in the FAs: simultaneously occurring with disease onset (most common) or sudden malignant change in longstanding benign FA. In one case series^14^ several disorders, including weight loss, fatigue, loss of appetite, and general deterioration, were found to suggest an immunological and endocrinological influence on the malignant transformation. Martin et al. had reported a lower progesterone and higher estradiol levels than normal in patients with FA of the breast.^15^ In addition, the authors also found a higher concentration of estrogen-binding sites in the FAs. Hormone receptor analysis is poorly reported globally in carcinomas arising from FAs.

In our study, 11 out of the 23 patients with breast carcinoma has strong estrogen receptor (ER+) positivity and one case of weak positivity. In a previous study, Rosen et al. had found 3 ER+ cases of FAs among 27 FAs.^16^ In addition, the authors also found that ER+ was more common in ILC than in IDC. Estrogenic influence on the highly sensitive terminal duct of the breast and highly concentrated ER+ receptors in receptor cells of FAs may initiate the malignant transformation.

The behavioral nature of carcinomas arising independently or from FAs are similar and thus treatment approach should be same. However, there is a debate pertaining the treatment approach for LCIS and DCIS which differs in their biological behaviors. DCIS within fibroadenoma is mostly managed by breast conservation surgery, whereas LCIS is usually managed by local excision and biopsy followed by periodic close surveillance. Despite the rare occurrence of this malignant transformation in benign FAs of the breast, surgeons must be aware of this rare clinical entity due to its multicentricity that arises either in the ipsilateral or contralateral breast.

## Conclusion

Here we report the clinicopathologic features of 23 cases of atypia and carcinomas arising within FAs of the breast diagnosed on CNB and the findings on the subsequent excision. In our study, we report an extremely low rate of breast carcinomas arising from FAs with important consideration of its clinical relevance for radiologists, pathologists and surgeons. Despite its low occurrence, FAs of the breast in older age group or with morphologic, morphologic heterogeneity on imaging should raise suspicion for malignant transformation and thus should be extensively evaluated with CNB and/or excisional biopsy. Due to the possible hormonal association of FAs with malignant transformation, blood hormonal analysis and receptor status assessment should be considered for selected patients. This hormonal influence should be studied in detail with a large population which may offer potential diagnostic, therapeutic, and prognosis guidance. A significant portion of cases of FA harboring atypia or malignancy diagnosed on CNB was upgraded on excision, suggesting that complete excision of these lesions should be the recommended method of clinical management.

## Data Availability

All data produced in the present work are contained in the manuscript

---

**Table 1:** All fibroadenoma cases with atypical/malignant lesions are numerically and in percent shown both on Core-Needle (CNB) and Excisional Biopsy.

| CNB Diagnosis (n/%) | Excisional Diagnosis (n/%) |
| --- | --- |
| LCIS (5/23) -22% | ☐ LCIS (2/5) – 40%<br>☐ LCIS + ALH (1/5) – 20%<br>☐ LCIS + DCIS (1/5) -20%<br>☐ LCIS + DCIS + ALH (1/5) – 20% |
| DCIS (4/23) – 17.4% | ☐ Invasive carcinoma with mixed ductal and lobular features (1/4) – 25%<br>☐ LCIS + DCIS (1/4) -25%<br>☐ LCIS + DCIS + IDC [classic and pleomorphic type] (1/4) – 25%<br>☐ No residual carcinoma (1/4) – 25% |
| ALH (7/23) - 30.5% | ☐ ALH (2/7)– 28.6%<br>☐ No residual ALH (2/7) -28.6%<br>☐ ALH+IDP (2/7) – 28.6%<br>☐ LCIS (1/7) – 14.2% |
| ADH (3/23) -13.1% | ☐ DCIS (1/3) – 33.3%<br>☐ Benign findings (2/3) - 66.7% |
| LCIS and ILC (1/23) - 4.5% | ☐ LCIS + DCIS (1) -100% |
| DCIS and IDC (2/23) - 9% | ☐ LCIS + DCIS (2/2) -100% |
| LCIS and ALH (1/23) – 4.5% | ☐ Benign findings [radial scars] (1/1) – 100% |

## Notes

### Competing Interest Statement

The authors have declared no competing interest.

### Funding Statement

This study did not receive any funding

### Author Declarations

All human participants' procedures were approved by the Institutional Review Board of Lenox Hill Hospital, Northwell Health, New York, USA.

## References

1. Carty NJ, Carter C, Rubin C, Ravichandran D, Royle GT, Taylor I. Management of fibroadenoma of the breast. Ann R Coll Surg Engl. 1995 Mar; 77(2): 127–130.

2. Love SM, Sue Gelman R, Silen W. Fibrocystic disease of the breast—a nondisease? N Engl J Med. 1982 Oct 14;307(16):1010–4.

3. Stafyla V, Kotsifopoulos N, Grigoriades K, Kassaras G, and Sakorafas GH: Lobular carcinoma in situ of the breast within a fibroadenoma. A case reports. Gynecol Oncol. 2004 Aug;94(2):572–4.

4. Diaz NM, Palmer JO, Mcdivitt RW. Carcinoma Arising Within Fibroadenomas of the Breast A Clinicopathologic Study of 105 Patients. Am J Clin Pathol. 1991 May;95(5):614–22.

5. Schiaffino S, Calabrese M, Melani EF, et al. Upgrade rate of percutaneously diagnosed pure atypical ductal hyperplasia: systematic review and meta-analysis of 6458 lesions. Radiology. 2020 Jan;294(1):76–86.

6. Murray MP, Luedtke C, Liberman L, Nehhozina T, Akram M, Brogi E. Classic lobular carcinoma in situ and atypical lobular hyperplasia at percutaneous breast core biopsy: outcomes of prospective excision. Cancer. 2013 Mar 1;119(5):1073–9.

7. L. Deschenes, S. Jacob, J. Fabia, A. Christen. Beware of breast fibroadenomas in middle-aged patients with women. Can J Surg. 1985 Jul;28(4):372–4.

8. Aydın OU, Soylu L, Ercan Aİ, Bilezikçi B, Koçak S. Invasive ductal carcinoma developing from fibroadenoma. J Breast Health. 2015 Oct; 11(4): 195–198.

9. Buzanowski-Konakry K, Harrison Jr EG, Payne WS. Lobular carcinoma arising in fibroadenoma of the breast. Cancer. 1975 Feb;35(2):450–6.

10. Wu YT, Chen ST, Chen CJ, et al. Breast cancer arising within fibroadenoma: collective analysis of case reports in the literature and hints on treatment policy. World J Surg Oncol. 2014 Nov 10;12:335.

11. Mcdvitt R. Breast carcinoma arising in solitaly fibroadenomas. Surg Gynecol Obstet. 1967;125:572–6.

12. Goldenberg VE, Wiegenstein L, Mottet NK. Florid breast fibroadenomas in patients taking hormonal oral contraceptives. Am J Clin Pathol. 1968 Jan 1;49(1):52–9.

13. Brown JM. Histological modification of fibroadenoma of the breast associated with oral hormonal contraceptives. Med J Aust. 1970 Feb 7;1(6):276–7.

14. Yoshida Y, Takaoka M, Fukumoto M. Carcinoma arising in fibroadenoma: case report and review of the world literature. Journal of surgical oncology. 1985 Jun;29(2):132–40.

15. Martin PM, Kuttenn F, Serment H, Mauvais-Jarvis P. Studies on clinical, hormonal and pathological correlations in breast fibroadenomas. J Steroid Biochem. 1978 Dec;9(12):1251–5.

16. Rosen PP, Menendez-Botet CJ, Nisselbaum JS, et al: Pathological review of breast lesions analyzed for estrogen receptor protein. Cancer Res 35:3187–3194, 1975.

